# Electronic data management for vaccine trials in low resource settings: Upgrades, scalability and impact of ODK

**DOI:** 10.1101/2021.02.08.20191908

**Authors:** Michael Marks, Sham Lal, Hannah Brindle, Pierre-Stéphane Gsell, Matthew MacGregor, Callum Stott, Martijn van de Rijdt, Guillermo Gutiérrez Almazor, Suman Golia, Conall Watson, Abdourahamane Diallo, Alhassane Toure, Catherine Houlihan, Patrick Keating, Hélène Martin, Ana-Maria Henao Restrepo, Yaw Anokwa, Chrissy h Roberts

## Abstract

**Background:** ODK provides software and standards that are popular solutions for off-grid electronic data collection and has substantial code overlap and interoperability with a number of related software products including CommCare, Enketo, Ona, SurveyCTO and KoBoToolbox. In combination with the use of statistical analysis software such as R, these tools provide fully open-source options for off-grid use in public health data collection, management, analysis and reporting. During the 2018-2020 Ebola epidemic in the North Kivu & Ituri regions of Democratic Republic of Congo, we leveraged ODK and other tools to support the DRC Ministère de la Santé RDC and World Health Organization in their efforts to administer an experimental vaccine (VSV-Zebov-GP) as part of their strategy to control the transmission of infection.

**Method:** New functions were developed to facilitate the use of ODK, Enketo and R in large scale data collection, aggregation, monitoring and near-real-time analysis during clinical research in health emergencies. We present open-source enhancements to ODK that include a built-in audit-trail, a framework and companion app for biometric registration of ISO/IEC 19794-2 fingerprint templates, enhanced performance features, better scalability for studies featuring millions of data form submissions, increased options for parallelization of research projects, and pipelines for automated management and analysis of data. We also developed novel encryption protocols for enhanced web-form security in Enketo.

**Results:** Against the backdrop of a complex and challenging epidemic response, our enhanced platform of open tools was used to collect and manage data from more than 280,000 eligible study participants who received VSV-Zebov-GP under informed consent. These data were used to determine whether the VSV-Zebov-GP was safe and effective and to guide daily field operations.

**Conclusions:** We present open-source developments that make electronic data management during clinical research and health emergencies more viable and robust. These developments will also enhance and expand the functionality of a diverse range of data collection platforms (Ona, KoBoToolbox etc.) that are based on the ODK software and standards.

**Funding:** This research is funded by the Department of Health and Social Care using UK Aid funding and is managed by the NIHR (PR-OD-1017-20001). The views expressed in this publication are those of the authors and not necessarily those of the Department of Health and Social Care.

## INTRODUCTION

ODK [1] provides open-source, community developed software and standards that have found broad utility in public health research [2, 3], epidemiology [4], disease mapping [5] and anthropological [6] studies in low-and-middle-income countries (LMICs). It has also been used to positive effect in clinical trials [7] as well as in disease surveillance [8] during outbreaks. The core ODK tools facilitate (1) electronic data collection (EDC) on Android devices though the ODK Collect App, (2) aggregation of the data on a web accessible ODK Aggregate server and (3) downstream management of data using ODK Briefcase a desktop software tool. ODK Briefcase has both a graphical user interface (GUI) and command line interface (CLI), the latter of which makes the off-server management of data automatable via command line tools. The core ODK tools are some of the most widely used EDC tools in the world, with over 400,000 active users each month and more than one million app installs to date. They also form the basis of the ODK ecosystem [9], which underpins the function of many other electronic data tools including Ona [10], KoBoToolbox [11], SurveyCTO [12], CommCare [13] and Enketo [14]. Application Program Interfaces (APIs) exist which can make ODK systems communicate with other common EDC tools including REDCap [15], DHIS2 [16], R [17] and others. The open-source Enketo system is an active participant in the ODK ecosystem and provides webform based data entry tools that harmonise well with ODK Aggregate to facilitate data collection either with Android devices (ODK Collect) or via a browser (Enketo).

At the time we began work on the current study, ODK Collect was able to perform asymmetric encryption on records, providing very high levels of security because once encrypted, no field operator or malicious actor in control of a device could decrypt or tamper with the data. Whilst Enketo provided highly desirable options for browser-based data entry, it was unable to perform encryption on records at the start of this project. Neither platform had capacity to perform audit actions in order to monitor enumerator behaviours during data entry & modification, whilst options for biometric registration of study participants were limited to a sophisticated but subscription-based fingerprint registration system offered by the not-for-profit SimPrints project [18].

In the wake of the 2013-2016 Ebola outbreak in West Africa, which affected around 28,000 people [19], the World Health Organization established the “R&D Blueprint” [20], bringing together stakeholders in research and development activities surrounding epidemics and health emergencies. The primary goal of the R&D Blueprint was to facilitate rapid deployment and evaluation of vaccines, tests and therapeutics that could be used to control epidemics and emergencies. This R&D Blueprint includes provisions to assess the safety and efficacy of experimental vaccines under expanded access (also known as ‘compassionate use’) programmes. Because of the experimental nature of unlicensed products, any expanded access must be assessed in the context of research.

The Democratic Republic of Congo (DRC) has experienced eleven documented outbreaks of Ebola virus [21–23] with the most recent three having occurred in May-July 2018 (Équateur Province), from July 2018 - July 2020 (North Kivu/Ituri Provinces) and from May-November 2020). The North Kivu epidemic is the second largest Ebola outbreak on record, with more than 3,296 cases and 2,196 deaths having been reported by late Dec 2019 [23]. Control efforts targeted against the infection were complicated by a number of factors which included regional conflict, high population density, community mistrust of the response and limited infrastructure for healthcare provision and communications in the affected areas.

During both the Équateur (2018) and North Kivu (2018-2020) outbreaks, the Ministère de la Santé RDC and World Health Organization attempted to use VSV-Zebov-GP [24], a live replicating candidate Ebola, to halt the epidemic. The vaccine was deployed using a ring vaccination strategy [25] wherein the contacts and ‘contacts-of-contacts’ of Ebola cases were traced and offered vaccination. Ring vaccination aims to halt the transmission of infection by providing a ring or “belt” of resistant individuals around cases of infection. The success of such approaches is highly dependent on good contact tracing and high coverage vaccination. In the context of the R&D that must accompany expanded access programmes, all participants in a ring vaccination study must be followed-up for some period (here at 30 minutes, 3 days and 21 days post-vaccination) to assess the safety of the product. Any cases of infection amongst vaccinees must also be linked to the vaccination data for efficacy estimates. In the face of such complexity, there is a significant need to collect, manage and analyse large amounts of data during a study such as this; particularly when the number of participants grows to hundreds of thousands and includes special/vulnerable groups such as pregnant women, infants and those with immune suppression.

When the Équateur outbreak was declared in May 2018, the partners of this study set out to develop the LSHTM Emergency and Epidemic Data Kit (EDK), a specialist implementation of ODK and other tools which encompassed an EDC, aggregation, analysis and monitoring system that (1) was scalable to potentially millions of data form submissions, (2) could work off-grid, for instance during long periods without internet connection, (3) was amenable to automation, (4) could facilitate near-real-time monitoring and data sharing, (5) was fully open-source, (6) had the capacity to register ISO/IEC 19794-2 fingerprints, (7) could be replicated in the case of further outbreaks or international spread and (8) could optionally generate an audit-trail for monitoring enumerator behaviours.

## METHOD

### Approach to platform development

We followed *Agile* principles of platform development, particularly with respect to (a) favouring the development of working software over comprehensive documentation, (b) involving our end-users & stakeholders in all stages and deferring negotiations over roles, responsibilities, contracture & funding, and (c) allowing teams to self-organise and adapt strategies in response to change. In practice we used tools that were familiar to non-experts including WhatsApp, Slack and GitHub to build a real-time development hub that allowed academics, clinicians, computer scientists, field-workers, WHO project-leads and ministry staff to communicate and contribute in real time to the development of the platform whilst working in several countries, multiple time-zones and hostile environments. During the early implementation phase, we operated a 24-hour working pattern, rotating work between staff in order to have a working platform in place within the first ten days of the outbreak and in time for the first vaccine doses to reach the field. Software developments to the ODK ecosystem were developed and integrated into the EDK system as and when they became available, with workarounds in place in the interim.

To ensure that all software developments became available to the widest possible user-base, we have implemented as many software changes as possible to the core ODK and Enketo systems, which is to say that the system we present should be considered a specialised deployment of tools which continue to be freely available through the parent projects ODK and Enketo. New features and standards added to ODK for the EDK system were reviewed by the ODK Technical Advisory Board [https://getodk.org/community/governance] and made available for comment on the community forum [https://forum.getodk.org/c/features]. The open availability of all the current developments of ODK contrasts with the approach taken by several beneficiaries of the ODK ecosystem including SurveyCTO, SimPrints & Ona; all of which control access to some components of their software.

### Conceptual Framework

The conceptual framework for the EDK system was (1) **an extensible project-oriented ODK server system** that could provide parallel server environments for multiple research studies (2) **software changes to the ODK ecosystem** which could facilitate efficient management of millions of submissions in near-real-time, (3) **strengthening the security of Enketo webforms** via form-level asymmetric encryption, (4) an **open-source biometrics framework** for registration of ISO/IEC 19794-2 fingerprints via low-cost consumer hardware, (5) **Audit trail features for ODK** and (6) **Automation of data management and analysis** using ODK Briefcase CLI, R, Rstudio, R Markdown & FlexDashboards. A schematic of the EDK system design is provided in Figure 1.

**Figure 1:**
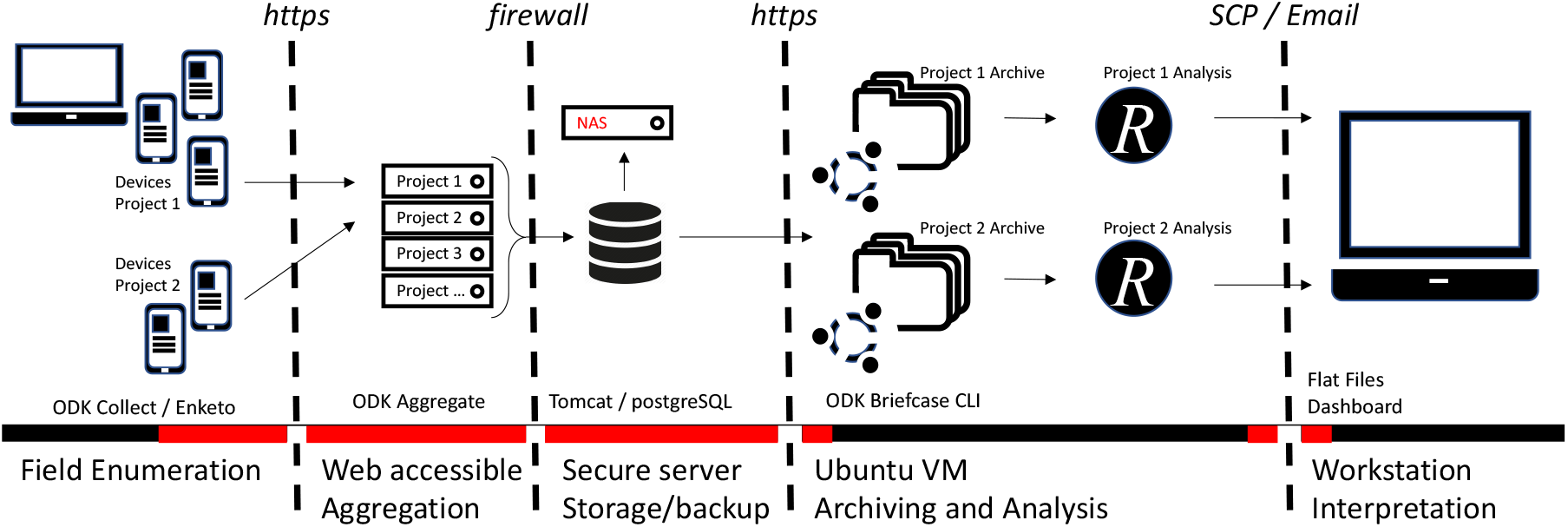
System design of the LSHTM Emergency and Epidemic Data Kit. Devices in the field are used to carry out data collection (enumeration) activities on browsers or an Android App. Those using Android have the option to register ISO/IEC 19794-2 fingerprint templates as part of the data collection. Encrypted records are submitted to one of many parallel web accessible ODK Aggregate server front-ends. All servers lead to a single PostgreSQL database and backup system. All backups contain only encrypted, non-human-readable form data. An Ubuntu virtual machine is scheduled to perform archiving (data pull, decrypt and export activities, including optionally analysis of fingerprint templates) and triggers analysis (data tidying, analysis and report generation) using R, R markdown and other open-source analytics tools. Outputs are securely copied or emailed to a workstation for end-user interactions. Areas highlighted in red show where data are stored or transferred in encrypted formats and are non-human readable.

### Project oriented ODK servers

In order to make the system extensible, for instance to make it possible to use an institutional installation of ODK servers for multiple research studies, we designed a server system infrastructure that allowed for the simultaneous operation of a number of parallel activities (projects). Each project was provisioned with its own dedicated ODK Aggregate server front end, unique URL and configurable user privileges. These project specific front-ends allowed for aggregation of data from EDC devices in the field, and for end-user level management of case report forms (CRFs) and individual project data entities in isolation from other projects, their CRFs and data. Behind the front-end, data from the many parallel projects were stored in a unified format on a single PostgreSQL database which was placed behind an institutional firewall and which was regularly backed up for data protection and recovery from failure. For each project, a data analysis pipeline was created on a virtual machine (VM) which was able to call data from the PostgreSQL database and to perform automated analysis, monitoring and reporting functions (Figure 1). Between-project and meta-analytics were also possible through this design. The addition of new projects required three steps including (1) the creation of a new ODK Aggregate front end, (2) design and deployment of project specific CRFs and (3) development of an analytics pipeline to match the needs of the project. An enhanced and improved approach to achieving a project-oriented design for ODK has now been adopted in ODK Central, a recently released ODK server which acts as a modern replacement for ODK Aggregate. Unlike ODK Aggregate, the ODK Central server has built-in project level management options that greatly simplify the handling of parallel projects.

### Software Performance Developments in the ODK ecosystem

ODK Briefcase is a desktop Java application which contains an application programming interface (API) that bridges the gap between study data on a server and the downstream analysis pipeline. It can both download individual data submissions (pull actions) from the ODK Aggregate server database and also parse, aggregate and export data to various formats, systems and backups. In the simplest terms, ODK Briefcase converts the many individual data files collected from the field into a single data set that is ready for analysis.

During the early development of EDK, we reached bottlenecks in the form of the time taken to perform pull and export actions. As the number of data submissions increased, so the time taken to process the data came to exceed the 24-hour analysis and reporting cycle of the field deployment. In order to make it possible to handle millions of CRF submissions to the EDK system without impacting significantly on time taken for pull and export operations, we introduced two performance related features to Briefcase, including “smart append” and “resume from last” controls.

The “smart append” feature speeds up exports of large datasets by remembering the full date and time of the last submission included in the most recent export for each form. By contrast to the historical approach (where all existing records were exported), the smart append feature exports only submissions which are new since the last export operation completed, appending these to the exported data from previous sessions.

The “resume from last” feature has an analogous function for the pull operations and speeds up downloads of submissions from ODK Aggregate (or ODK Central) by keeping track of information representing the last downloaded block of submissions and thereby only requesting new submissions in subsequent pulls. Previously, all submissions were always requested and Briefcase identified and discarded duplicates on download, leading to a potentially very large number of redundant network requests and database checks during each pull activity. The historical approach became prohibitively slow when the submission count reached the hundreds of thousands or more.

Both of these new features required storing new metadata in Briefcase and augmenting the graphical and command line interfaces. The use of metadata files to store the position was found to be preferable over storing this information in a system level preferences store as this change facilitated retention of the last pull/export positions in backups, thereby eliminating the need to start pulls and exports from the first submission after any system failure and/or recovery from backup. This work highlighted limitations of the Aggregate submission download API and has fed into the design of a replacement API.

### Strengthening security of Enketo webforms

Enketo is a suite of JavaScript tools which is a part of the ODK ecosystem and which among other uses can provide a web interface to ODK Aggregate servers. A longstanding feature of ODK Collect is its ability to asymmetrically protect CRF data at the level of the individual form using a powerful cryptographic process. This use of cryptography has particular value for research studies that collect sensitive data, such as those for which the EDK system was designed. This is especially valuable because study data encrypted at the level of the form are archived in the PostgreSQL database, in all web-facing servers and in all backups in an encrypted, non-human-readable format. Historically the Enketo system did provide functionality for secure end-to-end data transfers, but had no capability to encrypt CRF data at the form level.

The implementation of a Java-based encryption methodology in JavaScript was a challenging task because internet browsers have no native equivalent implementations of the algorithms used. We analyzed and documented the encryption methods used in ODK Collect, before reverse engineering them for use in Enketo. We then developed a robust process for asymmetric encryption in Enketo where the form data are encrypted using a random single-use symmetric encryption key, which is in turn then asymmetrically encrypted using a public Rivest–Shamir–Adleman (RSA) key that is inherited from the CRF. The resulting asymmetrically-encrypted symmetric encryption key is then passed to the server with the form submission and the form data can then be decrypted in ODK Briefcase using a private RSA key that is possessed only by authorised users. After testing with ODK Briefcase, we openly published an Enketo encryption algorithm that works across platforms and on all modern browsers. This implementation can handle and co-encrypt binary attachments, such as photos, movies and data from other sources including third party apps. We went on to author a sub-specification of the encryption algorithms, which has now been published as part of the open ODK XForms specification [9]. To facilitate the creation of an alternative ODK-compatible encryption/decryption library or application in the future, we separated our encryption implementation into its own module within the Enketo code-base [26].

### An open-source biometrics framework for ODK

To provide a basis for biometric registration of study participants, we developed an ODK Biometrics framework [27] of open-source tools for capturing (through the “Keppel” Android app and hardware scanner) and later matching (through a javascript CLI) ISO/IEC 19794-2 fingerprint templates. The Keppel app is a standalone project that is designed to interface with ODK Collect and its derivatives. The app currently works in combination with the ODK Collect app, which is able to call for delivery of fingerprint template data from the Keppel app using an Android “intent” (a software action which allows two apps to communicate with one another). The primary purpose of the Keppel biometrics framework is to (1) assist with the process of linking separate forms that relate to a specific study participant and (2) to confirm the identity of an individual seeking access to their study data as part of their rights of access. Keppel does not currently perform fingerprint matching processes on the Android device or mobile app. The Keppel CLI runs on Linux-like systems and is able to compare pairs of templates and to provide a score for the strength of the match between each pair. End-users are able to select thresholds for match/mismatch classification that provide the appropriate level of sensitivity and specificity for their work. The Keppel app currently works with the Mantra MFS100, a low cost (∼US$35) optical fingerprint scanner manufactured by Mantra Softech India PLC (https://www.mantratec.com/).

### Audit trail features for ODK

Many research studies and clinical trials require that enumerator behaviour during data collection can be fully audited by managers, external observers and regulators. We implemented a system in which ODK Collect is optionally able to generate a customisable log of enumerator behaviour and meta-data during data entry activities. If an ODK form is designed to include an audit, ODK Collect now creates a comma-separated-values (.CSV) audit file and appends data to this form as the form is opened or closed and as data are entered, changed or removed. The audit file is invisible to the end user during data collection and is encrypted using the standard ODK encryption protocols. The basic audit log file records a number of data entities, including *events, nodes, start*/*end timestamps, coordinates (lat/lon), old-value, new-value* and *current user* (Table 1). Events represent a particular user action such as opening a form, saving a form, or displaying a question. The audit system is able to optionally record the identity of the current user, to request the user’s identity each time the form is opened and also to log the current longitude and latitude of the device when data entry/modification took place. The *old-value* and *new-value* entities are used to record changes in *question* type events (i.e. changes made to the research data) and the system can optionally collect meta-data describing the reasons for changes having been made during a form editing session. Types of audit events are described in Table 2. The *nodes* audit entity describes the data field that was affected by the event and timestamps provide information on the time and duration of the event. Relying on the time reported by the device for timestamps could allow users or the network to change the device time and thereby manipulate the correctness of the audit log. For this reason, we only use device time for the form start timestamp. All subsequent event timestamps are therefore the result of elapsed time (which users cannot change) added to the form start timestamp. This means that whilst the timestamps themselves may potentially be inaccurate, the time elapsed within and between the timestamps are always accurate within one form editing session. Audit logs were implemented in ODK Collect v1.25.0. As with the biometrics framework, the audit trail feature is only available on the ODK Collect App and derivatives. No provision for audit in Enketo webforms is currently available.

**Table 1:**
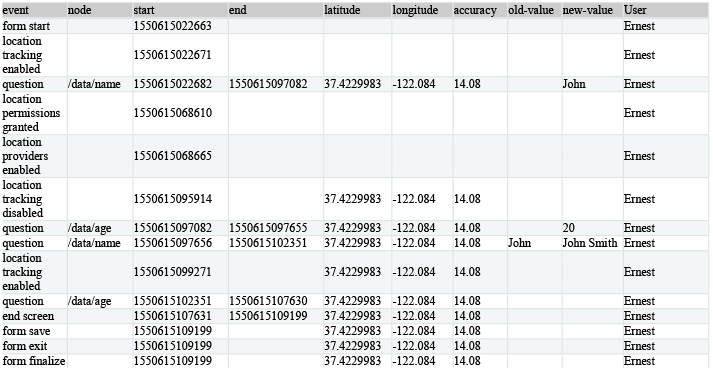
Exemplar ODK audit trail with coordinates, current user, timestamps and form events logged.

**Table 2:** Event types in ODK audit logs.

| Event | Description | Node? | Timestamps? | Coordinates? | Answers? |
| --- | --- | --- | --- | --- | --- |
| form start | Start filling in the form | No | **start** only | If on/available | No |
| question | View a question | Yes | Yes | If on/available | If enabled |
| group questions | View multiple questions on one screen (field-list) | Yes | Yes | If on/available | No |
| jump | View the jump screen | No | **start** only | If on/available | No |
| add repeat | Add a repeat | Yes | Yes | If on/available | No |
| delete repeat | Delete a repeat | Yes | Yes | If on/available | No |
| end screen | View the end screen | No | Yes | If on/available | No |
| form save | Save the form | No | **start** only | If on/available | No |
| form exit | Exit the form | No | **start** only | If on/available | No |
| form resume | Resume the form | No | **start** only | If on/available | No |
| form finalize | Finalize the form | No | **start** only | If on/available | No |
| save error | Error trying to save | No | **start** only | If on/available | No |
| finalize error | Error trying to finalize the form (probably encryption related) | No | **start** only | If on/available | No |
| constraint error | Constraint or required error on finalize | No | **start** only | If on/available | No |
| location tracking enabled/disabled | Toggle location tracking in Collect | No | Yes | If on/available | No |
| location providers enabled/disabled | Toggle location providers in Android | No | Yes | If on/available | No |
| location permissions granted / not granted | Toggle location permission in Android |  |  |  |  |

### Automation of data management and analysis

To enable an automated system to manage the pull, decrypt & export actions of Briefcase and to then perform data analysis and report generation steps, we set up an Ubuntu VM and scheduled automated operations using the *cron* utility. *Cron* is a powerful time-based job scheduler that is native to all Linux- and Unix-like systems. It allows computer code to be run on a regular basis and at predefined times. Cron requires very little computing experience and should be accessible to most users with support from an information technology team. On Windows systems it is possible to use the Windows Task Scheduler to achieve the same goal of automated pull, export and decrypt actions, or to use the recently released Windows Subsystem for Linux.

We conceptualised and implemented the data management tasks as two separate domains of work which included both *archiving* and *analysis* activities. *Archiving* consisted of the management of the raw form submissions, along with the maintenance of a set of up-to-date and human readable tables of raw data in comma separated value (CSV) format. The outputs of the archiving phase thereby represented the aggregated data from each CRF which formed the basis for all work in the *Analysis* phase. All activities in the *archiving* phase were automatically managed by ODK Briefcase. We used *cron* to run programmes (in the form of *bash* scripts) that were able to control ODK Briefcase via the CLI and to perform regular pull, export & decrypt actions. In order to protect the integrity of the data archive from human errors, we treated the CSV files in the archive as volatile entities that were subject to corruption if accessed by software other than ODK Briefcase. In order to ensure the integrity of the files, we isolated the archive from the analysis pipeline and used only copies of the CSV data files in downstream analysis.

The completion of a cron-scheduled archive process (pull, export, decrypt) triggered a series of R scripts using a Linux ‘pipe’ and leading to R’s native *Rscript* CLI command. On initiation of the R analysis, the first step was to make a working copy of the most up to date CSV files (from the archiving phase) in a system folder outside the ODK Briefcase managed *archive* folder.

The analysis of data included the use of both R and R markdown scripts, which eventually generated a large number of reports, charts, tables, line lists and other outcomes that had been conceptualised by the field and vaccination teams. We favoured the use of analysis tools that were both simple to use and openly available. We used primarily *ggplot2* [28], *plotly/ggplotly* [29], *leaflet* [30] and *flexdashboard* [31] to allow us to create interactive data visualisations that could be easily modified by future users with minimal need for coding. Because of the operational need to provide different teams with daily line-lists, a number of reports were automatically formatted as Microsoft Excel spreadsheets because Excel remains the default tool for many teams working with lists or tables. Figure 2 provides a schematic representation of some of the outputs of the system.

**Figure 2:**
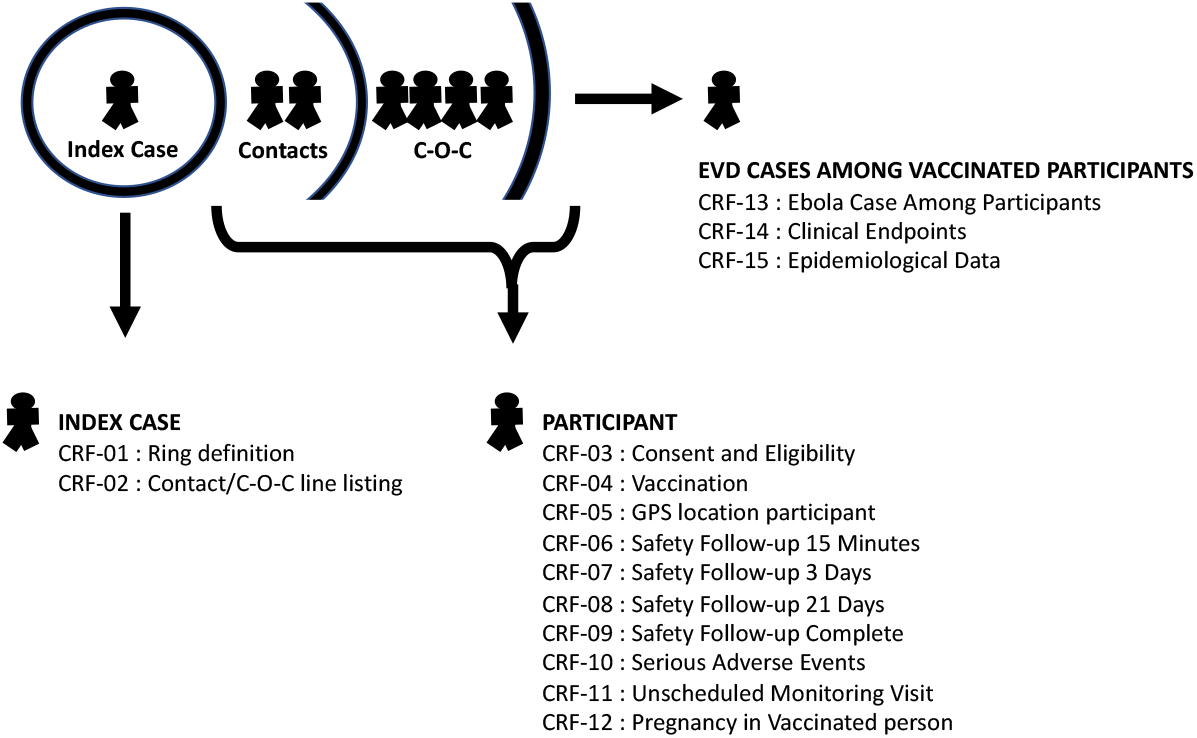
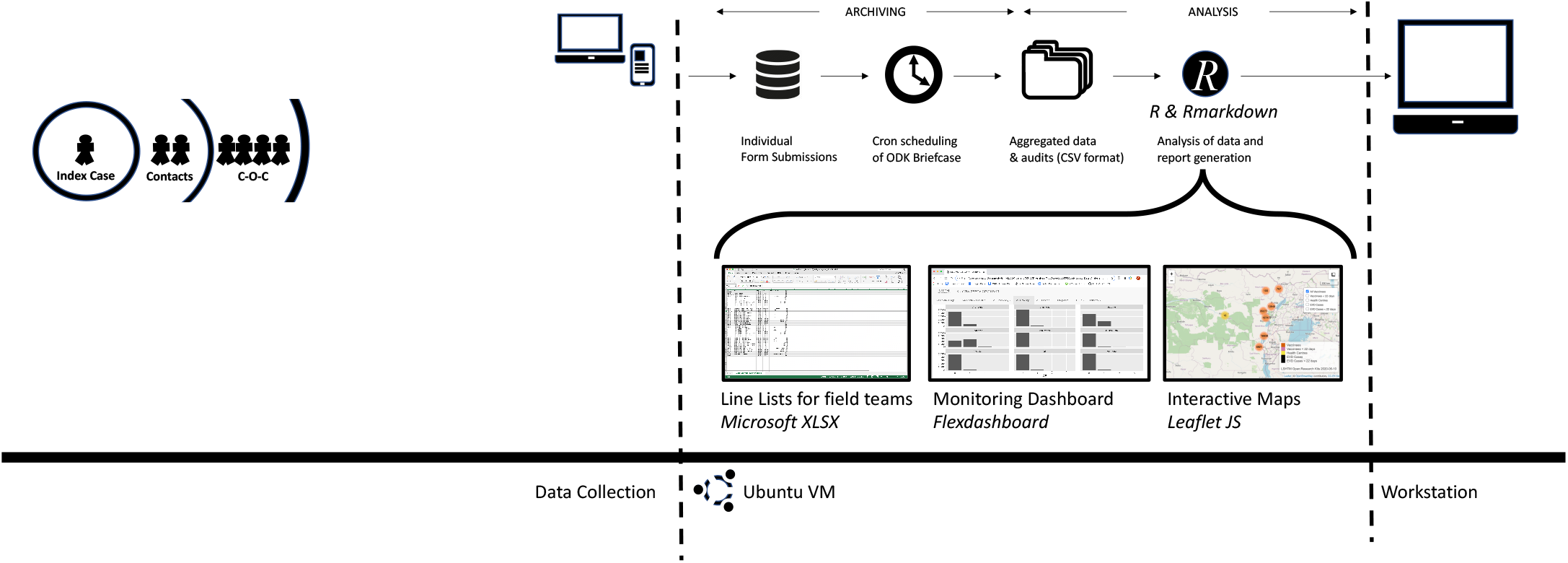
Fully customizable automated data flows. The time-based scheduler *cron* calls ODK Briefcase to perform data archiving and parsing of data from individual forms to aggregated tables of data in comma separated variable (CSV) format. *Cron* calls to R control Rmarkdown documents that perform statistical, geospatial and demographic analysis, along with data manipulation to create line lists, audit documentation, interactive monitoring dashboards (based on open tools such as Flexdashboard, LeafletJS etc), interactive maps and other outputs that are shared to workstations of partners in the field, senior academic team and internal/external monitors.

## RESULTS

We demonstrated the utility of a system for electronic data management during large-scale emergency clinical research settings through our case study of platform deployment during the response to the 2018-2020 North Kivu Ebola epidemic. Our need for near-real-time reporting during this work highlighted certain software behaviours that represented bottlenecks on the time taken for data management activities when using ODK. The large volumes of data that were being produced by the intense field operations of ring vaccination for Ebola virus control soon meant that the time taken for ODK Briefcase to download >500,000 form submissions from the server and then to export them to files for analysis was exceeding 24 hours. We solved this problem by implementing two new commands in Briefcase, firstly the ‘resume-from-last’ pull operation (ODK Briefcase v.1.14.0) and later the ‘smart-append’ export operation (ODK Briefcase v.1.17.0). Through the inclusion of a JavaScript Object Notation (.json) file within the app’s data storage directory, ODK Briefcase now stores the progress of the last *pull* and *export & decrypt* operations and since ODK Briefcase v.1.17.0 is not only able to resume from the last positions, but also to intelligently resume the position from restored backups and across mirrors or forks of the system. Between July 2018 and May 2020, data from more than 280,000 eligible study participants were recorded using the system. In our hands and when working with more than 1.75 million form submissions on the server, the time taken to perform a daily or *ad hoc* pull and export operation was reduced from hours-or-days to seconds-or-minutes, with the time taken now only dependent on the number of new submissions received since the last pull/export action and not on the cumulative number of submissions.

The system established an extensibility model by which the platform could be rapidly parallelised for use in other activities and projects. The effectiveness of this project-centred approach became clear in November 2018, when a small number of Ebola Virus transmission chains were traced into Uganda (which borders Eastern DRC) and where ring vaccination needed to start the next day. Whilst the design and delivery of ring vaccination activities undertaken in Uganda were identical to those in DRC, the work came under the jurisdiction of the Ugandan Ministry of Health and required separate administration and data management. We took advantage of our extensibility model to replicate the Ugandan ring vaccination system as a new ‘project’ and within one hour of the first reports of cases having been imported to the country, we had established a fully operational system dedicated to work in that country. In studies undertaken alongside the ring vaccination work, the VSV-ZEBOV-GP vaccine was also used in a programme of prophylactic vaccination studies that targeted healthcare workers (HCWs) and ‘front-line’ workers (FLWs) such as ambulance drivers, porters, burial teams and other people working in roles with high risk of exposure. These studies took place not only in DRC, but also in several neighbouring countries including South Sudan, Uganda, Burundi and Rwanda. Parallel EDK projects were used by teams led by each of the local ministries of health and the segregation of the project data between different jurisdictions had the additional benefit that we were able to comply with national and international data laws and best-practises, and also to ensure that each country had total authority over their own data. It was also possible to customise CRFs for local study requirements, as well as to change the language(s) used in the forms.

The international VSV-ZEBOV-GP prophylactic vaccination programme ultimately saw the vaccination of around 40,000 FLWs/HCWs in DRC (n ∼ 24,000), Burundi (n ∼4,000), Rwanda (n ∼3,500), South Sudan (n ∼3,000) and Uganda (n ∼7,000). In addition to our work during the North Kivu Ebola outbreak, we have also used the EDK system to provide data collection systems for research on other EVD research and in coronavirus disease 2019 (COVID-19) surveillance and vaccine/therapeutics trials; as well as in more than 200 non-emergency research projects. We have demonstrated the stability of the system by maintaining more than 100 parallel projects over a period of two years. The new audit trail and biometrics features became available towards the end of the VSV-ZEBOV-GP campaign and were not used in the field during that work. These features are however freely available to all users of ODK and have subsequently been used in research taking place during the COVID-19 pandemic.

## DISCUSSION

We present tools for automation of data management & reporting and open-source upgrades to the ODK ecosystem. These improvements implement data audit-trails, biometric participant registration, enhanced security for webforms and multiple performance upgrades that facilitate scaling and automation. These developments were driven by needs emerging in real time during the complex and challenging response to the North Kivu Ebola Virus Disease (EVD) epidemic. ODK and Enketo were selected for use in this work primarily because they provide the archetype for all ODK-related tools (Ona, KoboToolbox etc) and so developments made here affect functionality in a substantial proportion of modern EDC platforms and tools. ODK-based tools are also highly attractive to researchers and non-programmers because of the relative simplicity of CRF design in the ODK ecosystem (which is based around the use of Microsoft Excel spreadsheets). Further to this, ODK is amenable to scaling and has been used in some of the largest epidemiological studies ever undertaken, including the Global Trachoma Mapping Programme [5].

In order to maximise potential impact and accessibility to the outputs of this work, we ensured that all improvements made to ODK and Enketo were integrated with the open-source code-base of the main projects [26, 32]. Parties wishing to establish a functionally analogous system to the one described here are therefore encouraged to start with an installation of the most up to date releases of ODK’s core suite of tools. A large global community of ODK and Enketo users, along with users of related systems including Ona, KoBoToolbox, SurveyCTO and others will benefit equitably from these developments and these toolkits now represent a more complete data collection and management solution for robust clinical research studies.

The performance improvements made during our work on EDK have contributed to the design of a new, more performant ODK server system, ODK Central [32], which is designed to replace ODK Aggregate. ODK Central overcomes many of the performance limitations of Aggregate when submission counts get very large and also has native features that allow multiple projects to be managed from a single server environment; thereby removing the need to establish the more complex system of parallel server front-ends used in the work described above. The additional code we provide is sufficient to implement the biometrics framework [27] and to replicate the core infrastructure of an automated archiving and analysis system [34], although users are expected to provide their own CRFs and R scripts and we provide only exemplar CRFs and an analysis script which demonstrates the basic function of the automated analysis pipeline. The CRFs and analysis scripts used in vaccination activities surrounding the North Kivu epidemic contain sensitive materials and parties interested in these should contact the authors directly.

The use of webforms makes it possible to deploy ODK and other tools without the need for any software installation, or for any access to specific device types (i.e Android). As a result of our work, the Enketo system can now securely collect data using any device which has a modern browser such as Chrome, Firefox or Safari. The use of Enketo is therefore a simple means by which to increase the range of devices that can submit secure data to an ODK server to now include iOS devices, smart TVs, eReaders and both desktop and laptop PCs. Because they facilitate the collection of data through a simple URL, webforms are particularly useful when collecting data remotely in crowd-sourcing, electronic online surveys, clinics, laboratories and many other settings that are less well suited to the use of mobile apps. The novel JavaScript-based method of encryption that we have developed for Enketo is functionally analogous to the encryption system used by ODK Collect and was implemented in Enketo Express v1.72.0. The Enketo encryption system was recently deployed in an online survey that studied the effects of COVID-19 on health and wellbeing in ∼10,000 UK participants [33]. This system works both online and off-grid because Enketo can cache completed (encrypted) forms in the browser until an internet connection is found. Enketo forms are also a built-in feature of ODK Central, which further simplifies the work required to set up a system that includes secure web-forms as part of the data management solution.

Whilst the developments we present substantially improve the utility of ODK tools in public health research, this ecosystem is not without outstanding issues that represent barriers to more flexible use in this context. Primary amongst these is that data flow is unidirectional from devices towards the server and complex workflows are currently required to filter data back to the field via the server and back-end services. Options for synchronisation of data between devices and the server would greatly simplify the process of longitudinal study by making data from earlier time-points and activities more accessible to (and shareable between) enumerators in the field. The ability to open and edit, or to add to forms previously collected on a different device would also increase the range of tasks to which the tools could be applied and would reduce or remove the need to link different forms together in downstream analysis. A key challenge to implementation of both such capabilities comes from the need to be able to maintain options for both online and off-grid working, though requirements for off-grid capabilities are perhaps diminishing in many low and middle income countries as mobile connectivity becomes more accessible globally. Biometric frameworks for not only registering, but also recognizing study participants would strengthen data collection by confirming the identity of participants at key stages in data collection, in safety monitoring and in upholding rights of participant access and data security. Whilst SimPrints [18] already provides this type of functionality, no free-for-use biometric framework for ODK is currently available and future development of our open-source biometrics framework will seek to implement both on-device fingerprint matching/recognition and compatibility with a wider range of hardware devices. The ability to extensively view, manage, search, edit and audit changes to the PostgreSQL database from within the ODK Central server environment would further increase the range of applications of ODK tools in clinical research studies; as would tools for study randomisation. The combination of biometric registration, functional audit-trails and an auditable data management interface on the server would combine to make these open tools a more complete and attractive option for use clinical research; and in particular in GCP compliant clinical trials.

## Data Availability

Data sharing is not applicable to this article as no datasets were generated or analysed during the current study. All code and software developments described in this article are available from the repositories referenced in the manuscript section [Availability and Requirements]

https://github.com/getodk

https://github.com/LSHTM-ORK/ODK_Biometrics

https://github.com/LSHTM-ORK/EDK_Automation_Tools

## AVAILABILITY AND REQUIREMENTS

Project name: ODK

Project home page: https://getodk.org

Operating system(s): Collect: Android, Briefcase, Aggregate: any

Programming language: Java

Other requirements: e.g. Java

License: Apache 2.0

Any restrictions to use by non-academics: None

Project name: Enketo

Project home page: https://enketo.org

Operating system(s): Platform independent

Programming language: JavaScript, XSL, HTML, CSS

Other requirements: N/A

License: Apache 2.0

Any restrictions to use by non-academics: None

Project name: ODK Biometrics / Keppel

Project home page: https://github.com/LSHTM-ORK/ODK_Biometrics

Operating system(s): Android (App), Platform Independent (CLI)

Programming language: Kotlin, JavaScript

Other requirements: e.g.

License: MIT License

Any restrictions to use by non-academics: None

Project name: EDK Automation tools

Project home page: https://github.com/LSHTM-ORK/EDK_Automation_Tools

Operating system(s): Platform Independent (CLI)

Programming language: Bash, R

Other requirements: e.g.

License: MIT License

Any restrictions to use by non-academics: None

## LIST OF ABBREVIATIONS

APIs: Application Program Interfaces
CRFs: Case Report Forms
CSV: Comma-Separated-Values
CLI: Command Line Interface
COVID-19: Coronavirus Disease 2019
DRC: Democratic Republic of Congo
EVD: Ebola Virus Disease
EDC: Electronic Data Collection
EDK: Emergency and Epidemic Data Kit
FLWs: Front-Line Workers
GUI: Graphical User Interface
HCWs: Healthcare Workers
LMICs: Low and Middle Income Countries
RSA: Rivest–Shamir–Adleman
URL: Uniform Resource Locator
VM: Virtual Machine

## DECLARATIONS

### Ethics approval and consent to participate

The software platform developments required no specific ethics permission. Vaccination activities described in the case study were approved by the Democratic Republic of Congo (DRC) Ministry of health and DRC Public Health School ethics committee, or by the national research ethics committees of Rwanda, South Sudan, Uganda and Burundi. These activities were coordinated in collaboration with the World Health Organization and were carried out in accordance with GCP standards and with the Declaration of Helsinki. All vaccination activities were reviewed and monitored by an independent Data and Safety Monitoring Board (DSMB)

### Consent for publication

Not applicable

### Availability of data and materials

Data sharing is not applicable to this article as no datasets were generated or analysed during the current study. All code and software developments described in this article are available from the repositories referenced in the manuscript section entitled “Availability and Requirements”

### Authors’ contributions

MM & ChR : Conceptualised the study

MvdR, HB, CS, GGA, SL, MMcG, SG, YA & HM : Software/Hardware engineering & code

MvdR, GGA, CS, HM & YA : Software development on Enketo & ODK

CS : Software development on ODK biometrics framework

ChR & MM : Software development for automation and R analysis

PG, AHR, AD, AT, MM, SL, HB, CH, CW, PK & ChR : Platform development and field deployment of the Ebola data monitoring system

All authors : Wrote, reviewed and approved the manuscript

## Acknowledgements

We would like to express our profound appreciation and respect for the hundreds of field operatives who allowed us to support them with deployment of the EDK system during the Équateur (2018) and North Kivu (2018-2020) Ebola outbreaks. We are equally grateful to the people of North Kivu & Ituri and to our partners in the WHO teams and Ministries of Health of Uganda, Rwanda, Burundi and South Sudan. Thanks also to the members of the ODK community forum (https://forum.getodk.org/) and end users of the platform’s many non-emergency implementations. We would also like to acknowledge the invaluable administrative and financial management support provided by Esther Amon & Eleanor Martins.

## REFERENCES

1. ODK. https://getodk.org/. Accessed 16 Jun 2020.

2. Phiri TB, Kaunda-Khangamwa BN, Bauleni A, Chimuna T, Melody D, Kalengamaliro H, et al. Feasibility, acceptability and impact of integrating malaria rapid diagnostic tests and pre-referral rectal artesunate into the integrated community case management programme. A pilot study in Mchinji district, Malawi. Malar J. 2016;15:177.

3. Fornace KM, Surendra H, Abidin TR, Reyes R, Macalinao MLM, Stresman G, et al. Use of mobile technology-based participatory mapping approaches to geolocate health facility attendees for disease surveillance in low resource settings. Int J Health Geogr. 2018;17. doi:10.1186/s12942-0180141-0.

4. Rajput ZA, Mbugua S, Amadi D, Chepngeno V, Saleem JJ, Anokwa Y, et al. Evaluation of an Android-based mHealth system for population surveillance in developing countries. J Am Med Inform Assoc JAMIA. 2012;19:655–9.

5. Solomon AW, Pavluck AL, Courtright P, Aboe A, Adamu L, Alemayehu W, et al. The Global Trachoma Mapping Project: Methodology of a 34-Country Population-Based Study. Ophthalmic Epidemiol. 2015;22:214–25.

6. Dixon J, MacPherson E, Manyau S, Nayiga S, Khine Zaw Y, Kayendeke M, et al. The ‘Drug Bag’ method: lessons from anthropological studies of antibiotic use in Africa and South-East Asia. Glob Health Action. 2019;12:1639388.

7. Habtamu E, Wondie T, Aweke S, Tadesse Z, Zerihun M, Zewudie Z, et al. Posterior lamellar versus bilamellar tarsal rotation surgery for trachomatous trichiasis in Ethiopia: a randomised controlled trial. Lancet Glob Health. 2016. doi:10.1016/S2214-109X(15)00299-5.

8. Tom-Aba D, Olaleye A, Olayinka AT, Nguku P, Waziri N, Adewuyi P, et al. Innovative Technological Approach to Ebola Virus Disease Outbreak Response in Nigeria Using the Open Data Kit and Form Hub Technology. PloS One. 2015;10:e0131000.

9. ODK. ODK XForms Specification. https://getodk.github.io/xforms-spec/#encryption. Accessed 16 Jun 2020.

10. Ona. www.ona.io. Accessed 16 Jun 2020.

11. KoBo Toolbox. www.kobotoolbox.org. Accessed 16 Jun 2020.

12. Survey CTO. www.surveycto.com. Accessed 16 Jun 2020.

13. CommCare. https://www.commcarehq.org. Accessed 16 Jun 2020.

14. Enketo. https://enketo.org/. Accessed 16 Jun 2020.

15. REDCap_J: Research Electronic Data Capture. https://projectredcap.org/. Accessed 16 Jun 2020.

16. DHIS2. https://www.dhis2.org/. Accessed 16 Jun 2020.

17. R Core Team. R: A Language and Environment for Statistical Computing. R Foundation for Statistical Computing. 2014.

18. Storisteanu DML, Norman TL, Grigore A, Norman TL. Biometric fingerprint system to enable rapid and accurate identification of beneficiaries. Glob Health Sci Pract. 2015;3:135–7.

19. World Health Organisation. Ebola Situation Report - 30 March 2016 | Ebola. https://apps.who.int/ebola/current-situation/ebola-situation-report-30-march-2016. Accessed 16 Jun 2020.

20. World Health Organisation. An R&D Blueprint For Action To Prevent Epidemics. Plan of Action. 2016. https://www.who.int/blueprint/about/r_d_blueprint_plan_of_action.pdf. Accessed 16 Jun 2020.

21. Nsio J, Kapetshi J, Makiala S, Raymond F, Tshapenda G, Boucher N, et al. 2017 Outbreak of Ebola Virus Disease in Northern Democratic Republic of Congo. J Infect Dis. 2019.

22. Rosello A, Mossoko M, Flasche S, Van Hoek AJ, Mbala P, Camacho A, et al. Ebola virus disease in the Democratic Republic of the Congo, 1976-2014. eLife. 2015;4.

23. Aruna A, Mbala P, Minikulu L, Mukadi D, Bulemfu D, Edidi F, et al. Ebola Virus Disease Outbreak - Democratic Republic of the Congo, August 2018-November 2019. MMWR Morb Mortal Wkly Rep. 2019;68:1162–5.

24. Henao-Restrepo AM, Camacho A, Longini IM, Watson CH, Edmunds WJ, Egger M, et al. Efficacy and effectiveness of an rVSV-vectored vaccine in preventing Ebola virus disease: final results from the Guinea ring vaccination, open-label, cluster-randomised trial (Ebola Ça Suffit!). The Lancet. 2017;389:505–18.

25. Ebola ça Suffit Ring Vaccination Trial Consortium. The ring vaccination trial: a novel cluster randomised controlled trial design to evaluate vaccine efficacy and effectiveness during outbreaks, with special reference to Ebola. BMJ. 2015;351:h3740.

26. Enketo LLC. Official Enketo libraries and apps. https://github.com/enketo. Accessed 16 Jun 2020.

27. LSHTM Global Health Analytics Group. LSHTM-ORK/ODK_Biometrics. Kotlin. 2020. https://github.com/LSHTM-ORK/ODK_Biometrics. Accessed 16 Jun 2020.

28. Wickham H. ggplot2: Elegant Graphics for Data Analysis. New York: Springer-Verlag; 2009. doi:10.1007/978-0-387-98141-3.

29. Sievert C. Plotly for R. 2018. https://plotly-r.com.

30. Joe Cheng, Bhaskar Karambelkar, Yihui Xie. leaflet: Create Interactive Web Maps with the JavaScript “Leaflet.” 2019. https://CRAN.R-project.org/package=leaflet.

31. Richard Iannone, JJ Allaire, Barbara Borges. flexdashboard: R Markdown Format for Flexible Dashboards. 2018. https://CRAN.R-project.org/package=flexdashboard.

32. Nafundi. ODK code repository. https://github.com/getodk. Accessed 16 Jun 2020.

33. Nina Trivedy Rogers, Naomi Waterlow, Hannah E. Brindle, Luisa Enria, Rosalind M. Eggo, Shelley Lees, et al. Behavioural change towards reduced intensity physical activity is disproportionately prevalent among adults with serious health issues or self-perception of high risk during the UK COVID-19 lockdown. | medRxiv. doi:https://doi.org/10.1101/2020.05.12.20098921.

34. LSHTM Global Health Analytics Group. LSHTM-ORK/EDK_Automation_Tools. 2020. https://github.com/LSHTM-ORK/EDK_Automation_Tools. Accessed 16 Jun 2020.

